# Increased *Akkermansia* abundance is associated with increased colonic mucosal ω-3 fatty acids and decreased colonic mucosal PGE_2_ concentrations following healthy dietary pattern interventions

**DOI:** 10.1101/2021.10.11.21264748

**Authors:** S.B. Rifkin, A. Sen, D.K. Turgeon, R. Chan, M.T. Ruffin, D.E. Brenner, P.D. Schloss, Z. Djuric

**Affiliations:** Michigan Medicine Department of Internal Medicine, Ann Arbor, 48109, USA; Michigan Medicine Department of Family Medicine, Ann Arbor, 48109, USA; 10X Genomics, San Francisco, CA 94111; Penn State Cancer Institute, Hershey, PA, 17033, USA; Michigan Medicine Department of Immunology and Microbiology, Ann Arbor, MI, 48109,USA

**Author notes:** **Corresponding author:** Samara B Rifkin, MD, ScM, Division of Gastroenterology and Hepatology, University of Michigan School of Medicine.

**Keywords:** ω-3 polyunsaturated fatty acids (ω-3 PUFA), prostaglandin E_2_ (PGE_2_), cyclooxygenase (COX) -1 and COX-2, *Akkermansia*

## Abstract

Both increased dietary intake of ω-3 polyunsaturated fatty acids (PUFA) and subsequent increases in colonic mucosal ω-3 PUFA concentrations have been linked to improved gut barrier function and decreased risks of metabolic diseases and cancer. In addition, increased dietary ω-3 PUFA has been linked to eubiosis in mouse studies. Increased ω-3 PUFA function in part to reduce cyclooxygenase-(COX) mediated prostaglandin E_2_ (PGE_2_) production, a biomarker of cancer risk linked to compromised gut barrier function. We analyzed data from a dietary intervention study in individuals at increased risk of colon cancer to determine whether changes in the mucosal microbiome composition were associated with changes in colonic mucosal ω-3/ ω-6 PUFA ratio. Microbiome analyses of colonic biopsies before and after the dietary intervention from 86 participants were done by sequencing the V4 region of the 16S rRNA gene. Multivariable linear regression models were used to evaluate further whether changes in *Akkermansia* was associated with changes in each colonic tissue variable: ω-3/ω-6 PUFA ratios, PGE_2_ concentrations, and expression of COX-1 and COX-2. The median dietary intake and mucosal ω-3/ω-6 PUFA ratio increased after intervention. Greater increases in mucosal ω-3/ω-6 PUFA ratios after intervention were significantly associated with several changes in taxon abundance, including increased *Akkermansia muciniphilia* relative abundance. An increased abundance of *Akkermansia muciniphilia* also was associated significantly with decreased PGE_2_ concentrations but not with changes in COX expression. Further studies are warranted to elucidate mechanisms by which *Akkermansia* may affect or is affected by these pathways and the relative importance of individual dietary components.

## Introduction

Chronic low grade inflammation has been implicated in the rising prevalence of chronic diseases including obesity, diabetes, cardiovascular diseases and cancer.^1,2^ It has been suggested that Western diets containing relatively high saturated fatty acids and ω-6 polyunsaturated fatty acids (PUFA) but low monounsaturated fatty acids (MUFA) and ω-3 PUFA contribute to pro-inflammatory states and the rising prevalence of chronic diseases.^3^ In particular, consumption of ω-3 PUFA from fish and other foods results in a more favorable eicosanoid profile to reverse the chronic inflammation associated with reduced risks of obesity, diabetes and cancer.^4–6^ Dietary fats, however, have multi-faceted effects on inflammation and chronic diseases that also appears to include changes in the microbiome and gut barrier function.^2^

High saturated fat diets have been shown to compromise gut barrier function through mucus thinning and loss of junction barrier proteins leading to metabolic endotoxemia.^2,7,8^ Systemic exposures to bacterial lipopolysaccharides (LPS) are thought to mediate this process. LPS, a major component of the gram negative bacterial outer membrane, can be absorbed into the bloodstream to stimulate toll like receptor 4 (TLR4) with reactive elevations in serum pro-inflammatory eicosanoids and cytokines.^2,8,9^ Conversely, certain anti-inflammatory dietary components, including ω-3 PUFA, have been shown to prevent or reverse metabolic endotoxemia and chronic low grade inflammation by improving gut barrier function.^2,8,10^

Dietary ω-3 PUFA affect gut barrier function by several mechanisms. The ω-3 PUFA inhibit formation of pro-inflammatory eicosanoids such as prostaglandin E_2_ (PGE_2_) from the cyclooxygenase (COX-1) and COX-2-mediated oxidation of arachidonic acid (AA), and the reduction in PGE_2_ signaling reduces local inflammatory reactions to maintain normal barrier function.^11^ The ω-3 PUFA compete with AA as substrates for COX-1 and COX-2 enzymes and eicosapentaenoic acid ω-3 (EPA) also binds to COX-1 to allosterically inhibit AA oxygenation.^12^ In addition, ω-3 PUFA may prevent inflammation by influencing microbiome composition.^13^ A preponderance of studies have found that ω-3 PUFA administration increases LPS-suppressing bacteria including *Akkermansia mucinophila*.^2,10,14–19^ Increased tissue ω-3 PUFA in a transgenic mouse model that endogenously produces ω-3 PUFA led to lower levels of LPS-producing bacteria and higher levels of LPS-suppressing bacteria versus that in wild type mice.^10,14^ In human studies, the presence of *A. mucinophila* in the outer mucus layer of the colon was associated with reduction of metabolic endotoxemia and chronic low-grade inflammation brought on by high fat diet and obesity. These beneficial effects were attributed to the role of *A. mucinophila* in maintaining gut barrier function.^16^

Most studies aimed at investigating the relationship between PUFAs and intestinal microbiome composition have quantified the intestinal microbiota in excreted stool and measured PUFAs in the serum. Excreted stool, however, does not reflect the bacterial milieu adhering to the colonic mucosal surface.^20^ Bacteria adhering to the colonic mucin are in close proximity to the colon epithelial cells. Therefore, characterizing the bacteria associated with the colonic mucosa may be more meaningful to evaluate in the interaction between the diet and the gut microbiome. In the present study, we performed a secondary analysis of a study that recruited individuals with an elevated risk of colorectal cancer and randomized them to either a Healthy Eating diet or a Mediterranean diet. Both diet arms achieved similar increases in median dietary and median colonic mucosal ω-3/ω-6 PUFA ratios, but there was substantial inter-individual variability in fatty acid changes within each arm. This afforded the opportunity to evaluate whether changes in the colonic mucosal microbiome were associated with changes in colonic mucosal ω-3/ω-6 PUFA ratios. Because of its known role in promoting colonic integrity and limiting metabolic endotoxemia, we investigated further whether *Akkermansia* abundance was associated with colonic mucosal ω-3/ω-6 PUFA ratio as well as colonic mucosal COX-1 and 2 gene expression and PGE_2_ using multivariable linear regression models.

## Methods

### The Healthy Eating Study

Methods for conducting the Healthy Eating Study have been previously published.^21^ This study was conducted according to the Declaration of Helsinki and all procedures involving human subjects were approved by The University of Michigan Institutional Review Board (HUM00007622). This study was registered at ClinicalTrials.gov (registration no. NCT00475722). In brief, 120 persons at increased risk of colorectal cancer were enrolled from Ann Arbor, MI, and surrounding areas from July 2007 to November 2010. Increased risk was defined as having a family history of colorectal cancer in either one first-degree relative or two second-degree relatives, or having a personal history of colorectal adenoma or cancer. All study participants provided informed, signed consent prior to randomization.

A total of 94 participants completed 6 months of study. Participants underwent un-prepped flexible sigmoidoscopy before and after the 6-month intervention. Eight colorectal biopsy samples were obtained from the colon, 20 to 25 cm from the anal sphincter. Biopsy samples were flash frozen in liquid nitrogen within 5 seconds and kept at –70°C until analysis. Colon biopsy samples were available for microbiome analysis from 94 participants at baseline and from 86 participants after dietary intervention. Blood was collected after an overnight fast and prepared serum was kept frozen at -80 °C until analysis.

### Measures of fatty acids, prostaglandin E_2_, COX-1 and COX-2

Serum and colon biopsy samples were analyzed for complete fatty acid profiles as methyl esters using gas chromatography—mass spectroscopy (GC-MS). ω-3 PUFA was calculated by summing 18:3, 20:5, and 22:6, whereas ω-6 PUFA was calculated by summing 18:2 (n6), 20:3 (n6), 20:4 (n6). The ratio was then produced dividing ω-3 PUFA by ω-6 PUFA. PGE_2_ in colon mucosal biopsy samples was analyzed using liquid chromatography with mass spectrometry as previously described.^12^ COX-1 and COX-2 gene expression was quantified by Quantitative real-time PCR.^22^

### Dietary intervention

Participants were randomized to receive dietary counseling to follow either a Healthy Eating diet or a Mediterranean diet and the specific details of the two dietary interventions have been published previously.^21^ Overall, the change in dietary ω-3/ω-6 PUFA ratios and colonic mucosal ω-3/ω-6 PUFA ratios were similar between the two study arms, however there was inter-individual variation in fatty acid intakes and colonic tissue levels across the entire study population that allowed us to perform this secondary data analysis.^21^

Dietary data was obtained at baseline and 6 months using two 24-hour recalls and two days of written food records (four non-consecutive days per time point). The analysis of food records and recalls were performed with the Nutrition Data System for Research (2013 version; University of Minnesota, Nutrition Coordinating Center). Food and nutrient intakes were averaged over the four days of intake for each subject at each time point.

### Mucosal microbiome 16S rRNA sequencing and processing

DNA was isolated from biopsy samples using PowerMag Microbiome RNA/DNA Isolation Kit (Mo Bio Laboratories, Inc) and epMotion 5075 liquid handling system (Eppendorf). The hypervariable V4 region was amplified using primers described previously and16S rRNA gene sequences were processed using the mothur software package.^23^ FASTQ files were generated for paired end reads. Paired-end reads were merged into contigs, aligned to the SILVA 16S rRNA sequence database (version 138), and low quality sequences and chimeras were removed.^24^ Sequences were classified by training a naive Bayesian classifier with a 16S rRNA gene training set (Ribosomal Database Project (RDP)).^25^ Sequences were then clustered into operational taxonomic units (OTUs) using a 97 % similarity cutoff with the OptiClust clustering algorithm.^26^ Samples were rarefied to 1,047 sequences per sample, which eliminated ten of 179 samples.

### Statistical analysis

Individuals were categorized according to baseline colonic mucosal ω-3/ω-6 PUFA ratios tertiles. Baseline characteristics variables were tested for normality using the Shapiro-Wilk’s method in R (version 4.0.2), which did not follow a normal distribution. Therefore, these values were assessed using medians and interquartile ranges for continuous variables, and as frequencies and percentages for categorical variables. Additionally, fatty acid dietary intake, serum and colonic mucosal concentrations before and after dietary intervention did not follow a normal distribution. We therefore calculated median differences in these variables before and after intervention. Statistically significant changes in median differences in nutrient variables were assessed using Wilcoxon signed rank test.

We characterized variation in microbial community using metrics for 1) α diversity (Shannon index) to quantify intraindividual variation and 2) β-diversity-based distance metric (Bray-Curtis) to quantify the distance between individuals. These values were calculated using mothur software package. We compared Shannon index across serum, diet and colonic mucosal ω-3/ω-6 PUFA ratios tertiles using Kruskal-Wallis. Associations between microbiome variation and serum, diet and colonic mucosal ω-3/ω-6 PUFA ratios were assessed via permutational multivariate analysis of variance (PERMANOVA) both as continuous variables and using variables categorized into tertiles. To visualize the Bray-Curtis distance values between samples, Non-metric multidimensional scaling NMDS was calculated and results were plotted using the vegan package within the R program.

Linear regression was used to identify bacterial taxa whose relative abundance were associated with changes in of serum, diet and colonic mucosal ω-3/ω-6 PUFA ratios. The Benjamini-Hochberg (B-H) method was used to adjust P values in the multiple correlation analyses while controlling for the expected false discovery rate at 0.05. We then constructed a multivariable linear regression models for each colonic mucosal variable (ω-3/ω-6 PUFA ratio, PGE_2_ levels, COX-1 and COX-2 gene expression) to assess if a given colonic mucosal variable was associated with *Akkermansia* abundance. We modeled each colonic mucosal variable (ω-3/ω-6 PUFA ratio, PGE_2_ levels, COX-1 and COX-2 gene expression) at 6 months as a factor of log change in *Akkermansia* abundance levels after intervention, also adjusting for the colonic mucosal variable at baseline and age, sex and body mass index (BMI).

## Results

### Baseline characteristics of study participants

The 86 study participants with sequenced biopsy samples available at one or both time points were 74% female, had a median age of 54 years, and a median BMI of 27 kg/m^2^. A small proportion of participants (18%) were taking nonsteroidal anti-inflammatory drugs (NSAIDs). Baseline data are shown by tertiles of colonic mucosal ω-3/ω-6 PUFA ratios (Table 1). Age, BMI, sex, NSAID use, and PGE_2_ were not different across tertiles of colonic mucosal ω-3/ω-6 PUFA ratios.

**Table 1.** Baseline characteristics of Healthy Dietary Study participants by tertiles of colonic mucosal ω-3/ω-6 PUFA (polyunsaturated fatty acids) ratios at baseline

| Characteristic | Tertile 1, N = 28<sup>1</sup> | Tertile 2, N = 29<sup>1</sup> | Tertile 3, N = 31<sup>1</sup> | p-value<sup>2</sup> |
| --- | --- | --- | --- | --- |
| **$\omega$ -3/ $\omega$ -6 PUFA ratio tertiles** | **<0.07** | **0.07-0.27** | **>0.27** |  |
| Age (years) | 54 (48, 59) | 53 (49, 57) | 56 (48, 62) | 0.79 |
| Body Mass Index (kg/m<sup>2</sup>) | 26.1 (23.1, 29.5) | 27.2 (23.4, 32.0) | 26.9 (23.2, 28.8) | 0.32 |
| Female | 21 (75%) | 22 (76%) | 22 (71%) | 0.94 |
| Nonsteroidal anti-inflammatory drug (NSAID) use | 5 (18%) | 8 (28%) | 3 (9.7%) | 0.14 |
| Lipopolysaccharide (LPS) binding protein (μg/ml serum) | 15 (12, 24) | 15 (12, 19) | 15 (11, 21) | 0.91 |
| Prostaglandin E<sub>2</sub> (colon, pg/mg protein) | 16 (10, 26) | 12 (4, 23) | 14 (7, 24) | 0.38 |
| Shannon Index | 2.52 (2.39, 2.63) | 2.48 (2.37, 2.55) | 2.33 (2.02, 2.52) | 0.03 |
| Serum ω-3 PUFA<sup>2</sup> (%) | 3.16 (2.67, 3.62) | 3.38 (2.77, 3.73) | 4.33 (3.50, 5.33) | <0.001 |
| Serum ω-6 PUFA<sup>2</sup> (%) | 37 (35, 43) | 36 (33, 43) | 38 (33, 43) | 0.62 |
| Serum ω-3/ω-6 PUFA ratio | 0.08 (0.07, 0.10) | 0.09 (0.08, 0.10) | 0.11 (0.09, 0.14) | <0.001 |
| Dietary ω-3 PUFA (g/day) | 0.75 (0.58, 0.87) | 0.74 (0.56, 1.04) | 0.79 (0.56, 1.02) | 0.88 |
| Dietary ω-6 PUFA (g/day) | 6.94 (5.97, 8.14) | 6.75 (5.87, 8.38) | 6.20 (5.44, 7.47) | 0.32 |
| Dietary ω-3/ω-6 PUFA ratio | 0.11 (0.09, 0.13) | 0.11 (0.10, 0.14) | 0.13 (0.10, 0.16) | 0.24 |
| Colon tissue Saturated fatty acids<sup>2</sup> (%) | 32.1 (27.0, 34.8) | 32.6 (31.4, 35.3) | 32.6 (31.3, 33.1) | 0.21 |
| Colon tissue Monounsaturated fatty acids<sup>2</sup> (%) | 31.4 (28.1, 36.1) | 32.2 (28.7, 33.9) | 31.6 (29.1, 32.5) | 0.91 |

<sup>1</sup> Summary statistics are presented as median (IQR); n (%). The statistical tests performed were the Kruskal-Wallis test or the chi-square test of independence for categorical variables.
<sup>2</sup> Fatty acids in serum and colon tissue are shown as percent of total fatty acids analyzed.

### Effects of dietary intervention on fatty acid intake and concentrations

Median differences in fatty acid intake, serum and colonic mucosal concentrations after each dietary intervention are shown in Table 2. Dietary ω-3 PUFA increased in the Healthy Eating arm (P=0.04), while dietary ω-6 PUFA decreased in the Mediterranean arm (P<0.0001). The dietary ω-3/ω-6 PUFA ratio increased in both the Healthy Eating arm (P=0.01) and the Mediterranean arm (P=0.0002). Serum ω-3 PUFA increased in the Healthy Eating arm (P=0.004) and serum ω-6 PUFA decreased in the Mediterranean arm (P=0.04). The median serum ω-3/ω-6 PUFA ratio remained the same in the Healthy Eating arm and increased from 0.09 to 0.12 in the Mediterranean arm (P=0.03). Median colonic mucosal PGE_2_ concentrations did not change appreciably in the Healthy Eating arm (P=0.8) nor the Mediterranean arm after 6 months of intervention (P=0.7). The colonic mucosal ω-3/ω-6 PUFA ratio increased in both the Healthy Eating arm (P=0.03) and the Mediterranean arm (P=0.001). We confirmed that both arms achieved similar changes in colonic mucosal ω-3/ω-6 PUFA ratio. However, there was substantial inter-individual variation in colonic mucosal concentrations of all these analytes.

**Table 2.** Changes in dietary, serum and colonic mucosal fatty acid content before and after intervention in the Healthy Eating Study by diet arm assignment.

| Fatty acid % | Healthy Eating<br>(n = 42) | Mediterranean<br>(n = 44) | Total<br>(n = 86) |
| --- | --- | --- | --- |
|  | Median change<sup>1</sup> (95 CI%)<sup>2</sup> | Median change<sup>1</sup> (95 CI%)<sup>2</sup> | Median change<sup>1</sup> (95 CI%)<sup>2</sup> |
| MUFA<sup>3</sup> (%) (tissue) | 0.67 (-0.44, 1.63) | 0.36 (-1.10, 1.55) | 0.50 (-0.34, 1.25) |
| MUFA<sup>3</sup> (%) (serum) | 0.40 (-0.86, 1.52) | **1.77 (0.60, 3.13)** | **1.06 (0.21, 1.91)** |
| ω-6 PUFA<sup>4</sup> (%) (tissue) | -0.17 (-1.37, 0.88) | -0.29 (-1.22, 0.98) | -0.23 (-0.97, 0.54) |
| ω-3 PUFA<sup>4</sup> (%) (tissue) | **0.42 (0.07, 0.73)** | **0.48 (0.17, 0.76)** | **0.44 (0.22, 0.66)** |
| ω-3/ ω-6 ratio (tissue) | **0.01 (0.001, 0.02)** | **0.01 (0.006, 0.03)** | **0.01 (0.007, 0.02)** |
| ω-6 PUFA<sup>4</sup> (%) (diet) | -0.20 (-0.91, 0.62) | **-1.81 (- 2.50, -1.15)** | **-1.04 (-0.54, -1.54)** |
| ω-3 PUFA<sup>4</sup> (%) (diet) | **0.11 (0.005, 0.24)** | 0.07 (-0.11, 0.26) | 0.08 (-0.007, 0.20) |
| ω-3/ω-6 ratio (diet) | **0.02 (0.004, 0.03)** | **0.05 (0.02, 0.09)** | **0.03 (0.02, 0.05)** |
| ω-6 PUFA<sup>4</sup> (% , serum) | -0.13 (-1.26, 1.01) | **-1.73 (-2.92, -0.41)** | **-1.04 (-1.73, -0.25)** |
| ω-3 PUFA<sup>4</sup> (% , serum) | **0.26 (0.02, 0.67)** | 0.34 (-0.06, 0.71) | **0.32 (0.10, 0.57)** |
| ω-3/ω-6 ratio (serum) | 0.01 (-0.001, 0.02) | **0.01 (0.002, 0.02)** | **0.01 (0.003, 0.02)** |
| <sup>1</sup>Values are the change in mean percentages for each fatty acid.<br><sup>2</sup> 95% confidence intervals calculated with Wilcoxon signed rank test.<br><sup>3</sup> Monounsaturated fatty acid (MUFA)<br><sup>4</sup> Polyunsaturated fatty acid (PUFA)<br>Boded values are significant with P-value<0.05 |  |  |  |

### Effects of fatty acid concentrations on gut microbiota

To test the hypothesis that fatty acids can influence the microbiome, we examined how global microbiome features, including within community variation (alpha diversity with Shannon index) and between community variation (beta diversity using Bray-Curtis distance), varied according to colonic mucosal, serum and dietary fatty acid levels.^22^ The Shannon index was inversely associated with baseline colonic mucosal ω-3/ω-6 PUFA (P=0.02), but not associated with serum ω-3/ω-6 PUFA ratio at baseline nor any of these variables at 6 months after intervention (Figure 1).

**Figure 1.**
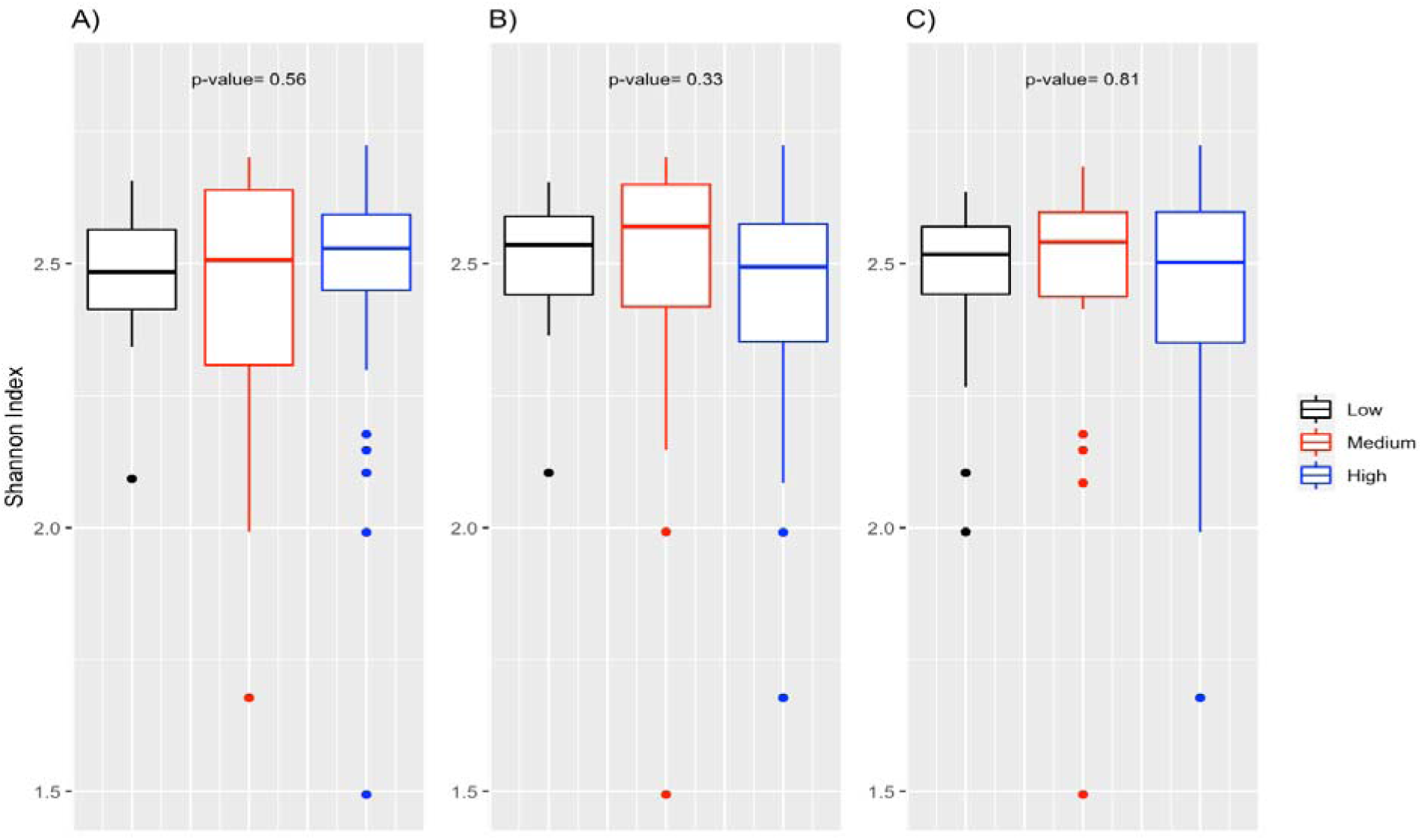
Alpha diversity, accessed by Shannon Index, across tertiles of A) dietary, B) serum and C) colonic mucosal ω-3/ω-6 PUFA levels after 6 months of dietary intervention.

When we performed PERMANOVA analysis, we found no differences in beta diversity or community distance according to diet (P= 0.5), or colonic mucosal ω-3/ω-6 PUFA ratio at 6 months (P= 0.1), however serum ω-3/ω-6 PUFA ratio at 6 months showed a stronger association with beta diversity (P=0.003). We also performed PERMANOVA analysis using dietary (P=1.00), serum (P=0.09) and colonic mucosal ω-3/ω-6 PUFA ratio (P= 0.46) categorized into tertiles and found no differences in beta diversity (see Figure 2). We found that when we performed pairwise PERMANOVA on serum ω-3/ω-6 PUFA ratio levels tertiles at 6 months that none of the differences were significant (P=0.5).

**Figure 2.**
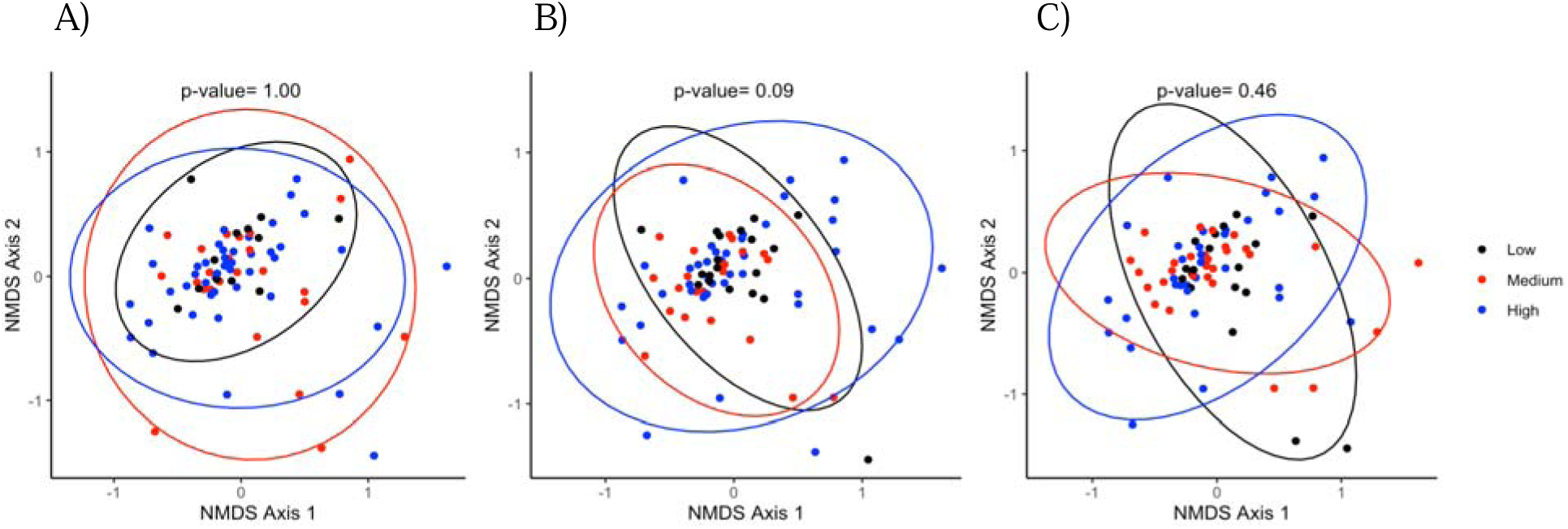
Between microbial community differences across tertiles of A) dietary, B) serum and C) colonic mucosal ω-3/ω-6 PUFA levels following dietary intervention. The circles represent the different tertiles of ω-3/ω-6 PUFA ratios in diet, serum and colon biopsies. Non-metric multidimensional scaling (NMDS).

### Abundance of taxa by colonic mucosal, serum and dietary ω-3/ω-6 PUFA ratios at baseline

We identified several differentially abundant genera that were positively associated with baseline colonic mucosal ω-3/ω-6 PUFA ratio including *Varibaculum, Rhizobium, Porphyrmonas, Mogibacterium, Megamonas, Coprobacillus, Butyrcicoccus, Actinomyces* and unclassified members of *Erysipelotrichaceae*. Unclassifed members of *Lachnospiracea* were negatively associated with ω-3/ω-6 PUFA ratio (Figure 3).

**Figure 3.**
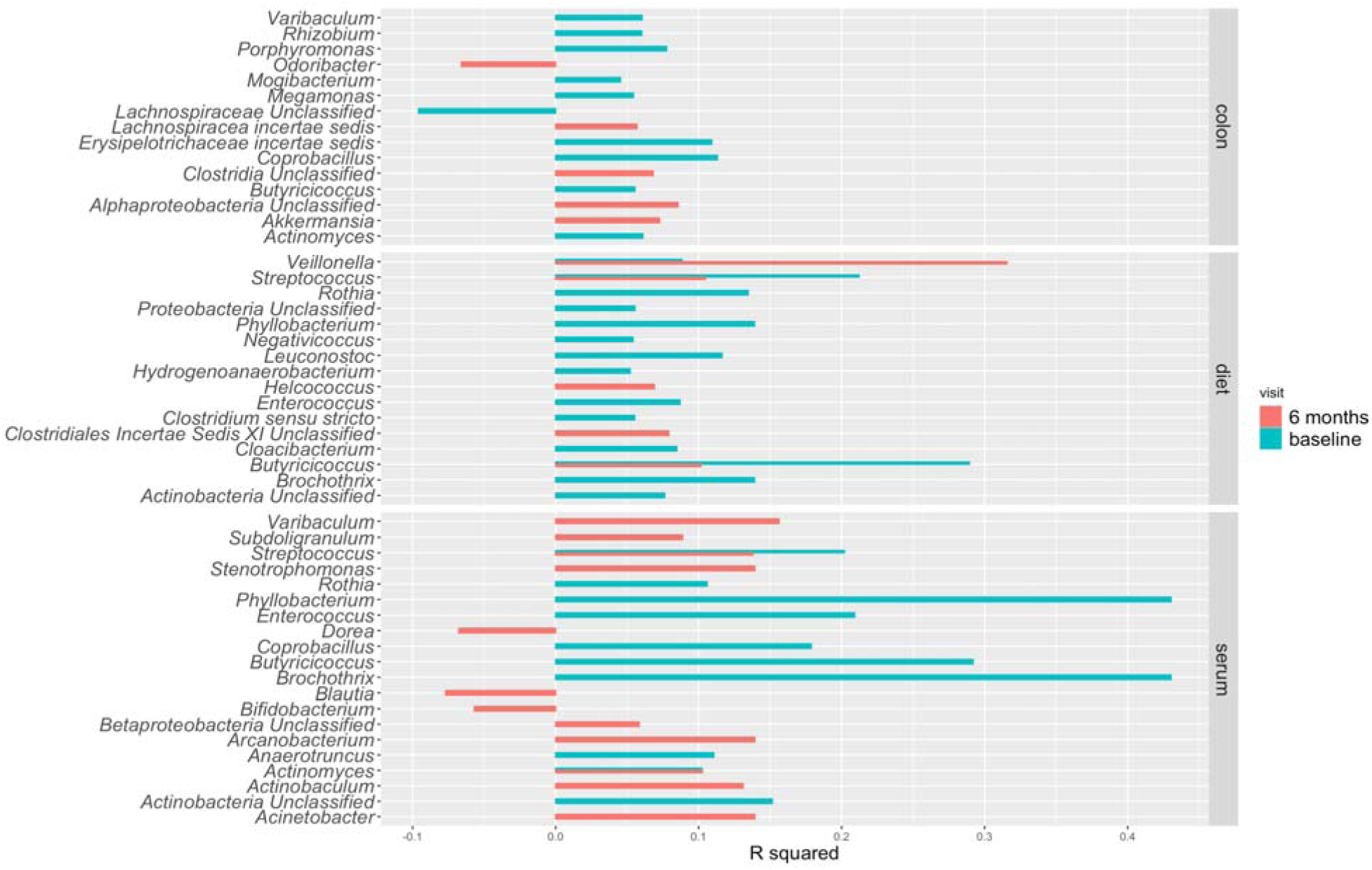
Correlation between taxon abundance and dietary, serum and colonic mucosal ω-3/ω-6 PUFA ratios before and after 6 months of dietary intervention.

The following genera were positively associated with baseline serum ω-3/ω-6 PUFA ratio (Figure 3): *Rothia, Phyllobacterium, Negativicoccus, Leuconostoc, Hydrogenoanaerobacterium, Enterococcus, Clostridium sensu stricto, Cloacibacterium, Brochothrix* and unclassified members of *Proteobacteria* and *Actinobacteria. Rothia, Phyllobacterium, Enterococcus, Coprobacilus, Butyrcicoccus, Brochothrix, Anaeotruncus* and unclassified member of *Actinobacteria* were positively associated with dietary ω-3/ω-6 PUFA ratio levels.

### Abundance of taxa by colonic mucosal, serum and dietary ω-3/ω-6 PUFA ratios after 6 months

We identified several differentially abundant genera whose relative abundance were positively associated with colonic mucosal ω-3/ω-6 PUFA ratio including unclassified members of *Alphaproteobacteria, Clostridia* and *Lachnospiracea*, and *Akkermansia* at 6 months. *Odoribacter* was negatively associated with colonic mucosal ω-3/ω-6 PUFA ratio at 6 months (Figure 3).

The following genera were positively associated with serum ω-3/ω-6 PUFA ratio (Figure 3): *Varibaculum, Acinetobacter, Arcanobacterium, Stentotrophomonas, Streptococcus, Actinobaculum, Actinomyces, Subdoligranulum*, and unclassified members of *Betaproteobacteria*, while *Blautia, Dorea* and *Bifidobacterium* were negatively associated at 6 months. *Veillonella, Streptococcus, Butyricicoccus, Helococcus* and unclassified members of *Clostridiales* were positively associated with dietary ω-3/ω-6 PUFA ratio levels.

### *Akkermansia* abundance and colonic mucosal ω-3/ω-6 PUFA ratio, COX gene expression, PGE_2_ concentration

We further investigated whether *Akkermansia* abundance was associated with colonic mucosal ω-3/ω-6 PUFA ratio as well as colonic mucosal COX-1 and 2 gene expression, PGE_2_ levels adjusting for age, BMI, sex and dietary ω-3/ω-6 PUFA ratio intake. COX-1 and COX-2 were not associated with ω-3/ω-6 PUFA ratio. We investigated whether these colonic mucosal nutrient concentrations, gene expression and metabolite concentrations were associated with levels of *Akkermansia* abundance using two different linear models.

To assess the effect of change in *Akkermansia* abundance on colonic mucosal ω-3/ω-6 PUFA ratio, PGE_2_, COX-1 and COX-2 gene expression following the dietary intervention, we modeled each colonic mucosal variable at 6 months as the dependent variable adjusting for each colonic mucosal variable at baseline and change in *Akkermansia* abundance using linear regression (Table 3). We found that a positive log change in *Akkermansia* following the dietary intervention was significantly associated with increased colonic mucosal ω-3/ω-6 PUFA ratios at 6 months after adjusting for age, sex, BMI, and dietary ω-3/ω-6 PUFA ratios (P=0.02). When we modeled colonic mucosal PGE_2_ at 6 months with adjustment for baseline values, a negative log change in *Akkermansia* following the intervention conversely was associated with a positive change in colonic mucosal PGE_2_ (P=0.01). COX-1 or COX-2 expression at 6 months were not associated with log change in *Akkermansia* abundance (P=0.3) and (P=0.5), respectively.

**Table 3.**
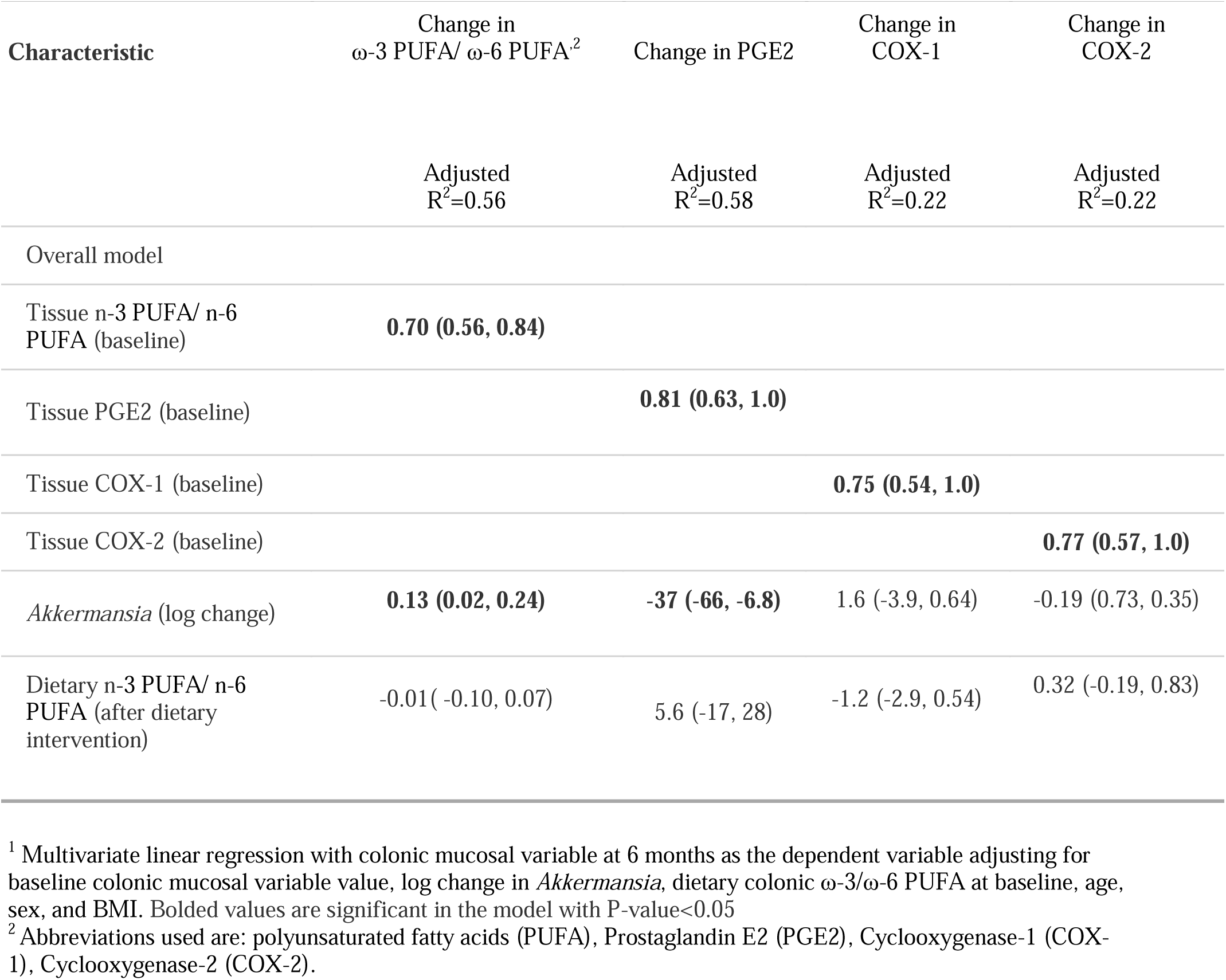
Linear regression modeling of colonic mucosal variables (ω-3/ω-6 fatty acid ratios, PGE_2_ concentration in colon, cyclooxygenase (COX)-1 and 2 gene expression in colon) at 6 months as a factor of log change in *Akkermansia* abundance levels in all 86 individuals after dietary intervention adjusting for baseline colonic mucosal variable values. The data shown is the adjusted f3 regression coefficient and 95% confidence interval (CI). The adjusted R^2^ represents the fraction of variability in the colonic mucosal variable at 6 months that is explained by the linear regression model^1^.

## Discussion

The two dietary intervention arms resulted in similar changes in serum and colonic mucosal ω-3/ω-6 PUFA ratios (Table 2). We therefore analyzed changes in colonic mucosal measures and in bacterial populations in all 86 study participants combined. The colonic mucosal ω-3/ω-6 PUFA ratio increased in the combined study population. Overall, we found that differences in PUFA incorporated in colonic tissues were not associated with global changes in the microbiome, but they were associated with specific microbiome changes. Alpha diversity, i.e. within community diversity, was associated with colonic mucosal ω-3/ω-6 PUFA ratios at baseline, but otherwise not associated with dietary or serum ω-3/ω-6 PUFA ratio at baseline or any nutrient ratios at 6 months (Figure 1). Beta diversity, i.e. inter-sample diversity, varied according to serum but not colonic mucosal ω-3/ω-6 PUFA concentration at 6 months (Figure 2). Our results analyzing differential taxon abundance associated with changes in ω-3/ω-6 PUFA ratio are consistent with both mouse studies and clinical interventions, which also found that higher ω-3 PUFA levels may influence the abundance of short chain fatty acid (SCFA) producing bacteria.^10,15,17– 19,27,28^ Several studies involving ω-3 supplementation reported increases in SCFA-producing bacteria.^15,17,19,29,30^ We also found a positive association between *Lachnospiracea* and colonic mucosal ω-3/ω-6 PUFA ratios, but an inverse association with *Odoribacter*, both producers of SCFA (Figure 3).

In subsequent analyses, we focused on the association between *Akkermansia* abundance and colonic mucosal ω-3/ω-6 PUFA ratio concentrations that had emerged in taxon-wide analyses since a lot of information is available on the role of *Akkermansia muciniphilia* in colon health.^1,8,16^ In addition to colonic mucosal PUFA, we explored the association between *Akkermansia* abundance and PGE_2,_ a pro-inflammatory mediator in the colon.^31^ PGE_2_ is produced from COX-mediated oxygenation of arachidonic acid, an ω-6 PUFA. We found that increased abundance of the *A. muciniphilia* after 6 months of dietary intervention was significantly and independently associated with increased colonic mucosal ω-3/ω-6 PUFA ratios and with decreased PGE_2_ concentrations using two different types of linear models, adjusting for age, sex and BMI. This did not appear to be due to changes in COX-1 or COX-2 expression in colonic biopsy tissues.

*A. muciniphilia* is found within the mucin layer and therefore is likely to impact the gut barrier function. Higher levels of *Akkermansia* abundance have been linked to decreased serum LPS levels, improved insulin resistance and low grade chronic inflammation^.16^ While obesity and high fat diets were associated with decreased levels of *Akkermansia* abundance, *Akkermansia* supplementation improved metabolic endotoxemia, hyperglycemia, and adiposity in mice fed a high fat diet.^16^ Additionally, *Akkermansia* is enriched in people who consume other diets that are known to enhance barrier function and improve metabolic endotoxemia, including general anti-inflammatory dietary patterns and functional foods rich in polyphenols including extracts of black raspberry, concord grape and cranberry.^2,32–36^

Our findings of increased *Akkermansia* levels with higher colonic tissue ω-3/ω-6 PUFA ratios are compatible with prior animal studies and clinical interventions.^9,14,17,18^ Several murine models have also demonstrated that fish oil supplementation resulting in increased colonic mucosal ω-3/ω-6 ratios led to higher *Akkermansia* abundance levels, among other taxon.^9,14,18^ The association between *Akkermansia* and two variables in the same pathway in the expected directions makes a stronger argument for a causal relationship, however we did not find an association between increased ω-3/ω-6 PUFA ratios and decreased PGE_2_ concentrations following the dietary intervention. Furthermore, the association between PGE_2_ and *Akkermansia* abundance was not weakened when ω-3/ω-6 PUFA ratio was included in the model.

While there is accumulating evidence that colonic ω-3 PUFA may modify the colonic mileau and encourage the growth of *Akkermansia*, it is unclear how *Akkermansia* abundance may be influencing colonic mucosal PGE2 levels or vice versa. A relevant mouse study demonstrated that increased SCFA levels promoted an increased PGE_1_/PGE_2_ ratio that in turn stimulated myofibroblast to express MUC-2, an important component of barrier integrity.^37^ Conversely, another mouse study found that increased PGE_2_ through its receptor EP4 reduced SCFA-producing bacteria leading to repression of intestinal Treg accumulation and accumulation of intestinal inflammation.^38^ While we speculate that colonic mucosal PUFA may influence the mucosal adherent microbiome, but the reverse or both directions could be true. Furthermore, we hypothesize that *Akkermansia* abundance may lead to improved gut barrier function and limit PGE_2_ production, but the reverse could be true. Therefore, the direction of the proposed effect cannot be determined from a statistical model and should also be interpreted with caution.

Both increased ω-3 PUFA and *A. muciniphilia* may alleviate the pathogenesis of obesity and high saturated fat diet on chronic low grade inflammation by limiting metabolic endotoxemia and strengthening gut barrier function.^16,39^ In transgenic fat-1 mice that endogenously convert ω-6 PUFA to ω-3 PUFA, transgenic fat-1 mice harbored lower levels of LPS producing bacteria and higher levels of LPS-suppressing bacteria (*Bifidobacterium, A. muciniphilia*) as compared with wild type mice.^14^ These mice also demonstrated increased expression of intestinal alkaline phosphatase (IAP), an anti-inflammatory enzyme located on the brush border that protects against pathogenic bacteria. Notably IAP promotes anti-inflammatory effects on the GI mucosa by dephosphorylating microbial LPS and microbiota-produced ATP, an important inflammatory mediator.^14^ IAP has also been linked to improved gut barrier function and has been shown to alleviate metabolic disease.^14,40^ The authors hypothesized that increased tissue ratio of ω-3/ω-6 PUFA ratios led to increased expression of IAP which cultivated an intestinal milieu favoring growth of LPS-suppressing bacteria.^14^

One strength of this study is the availability of colonic tissue for the fatty acid measures. Few studies have assessed the effect of colonic mucosal ω-3 PUFA on microbiome, and most studies have relied on self-reported diet, serum or RBC concentrations of ω-3 PUFA. Fatty acids are absorbed efficiently by the proximal small intestine with little residual traveling to the colon to affect the luminal microbiome composition.^41^ Therefore, the PUFA composition of colonic tissue more likely influences microbiome composition via indirect effects on the nature of the colonic barrier. The beneficial effects of colonic mucosal ω-3 PUFA have been shown to be manifested in part due to improved gut barrier function in both animal models and human intervention studies.^39,42^ In addition, our study also sequenced the microbiome adhering to colonic mucosal biopsies. Colonic mucosal fatty acids are more likely to affect bacteria associated with the mucosa, such as *A. muciniphilia*, a bacterium known to reside in the mucin in close proximity to the mucosa.

There are also several limitations to this study that should be recognized. The data in this study was analyzed using 16S rRNA and therefore our ability to determine changes in microbial function is limited. In addition, 16S rRNA analysis only allows resolution to the genus level. However, *A. muciniphilia* is the only bacterium of its genus known to colonize humans and therefore misclassification for this finding is unlikely. Lastly, sample size limited our ability to adjust for multiple covariates simultaneously.

In summary, we report a positive association between mucosal *Akkermansia* abundance and colonic tissue ω-3/ω-6 PUFA ratios and a negative association with colonic PGE_2_ concentrations in subjects who had undergone dietary intervention, either Mediterranean or Healthy Eating, both of which resulted in similar dietary changes with the exception of monounsaturated fats. These results are consistent with animal models and human studies that provide accumulating evidence of a connection between ω-3 PUFA and increased *Akkermansia* abundance on gut barrier function. Our growing understanding of how both ω-3 PUFA and *Akkermansia* improve gut barrier function and potentially limit metabolic endotoxemia is compelling as a causal association between diet, microbiome and chronic low-grade inflammation. Future studies exploring the effect of colonic mucosal ω-3/ω-6 PUFA ratios on the microbiome, on *A. muciniphilia* in particular, and gut barrier function are warranted.

## Data Availability

Microbiome sequences are available on SRA through NCBI.

## Acknowledgements

We thank all the individuals who volunteered to participate in the Healthy Eating Study. We acknowledge the contributions of Maria Cornellier in conducting the dietary counseling, Mary Rapai for study recruitment and coordination, Dr. Elkhansa Sidahmed for carrying out the gene expression analyses, Faith Umoh for carrying out the assays for LBP, and Jianwei Ren for the analyses of fatty acids, carotenoids and eicosanoids.

## Authors’ contributions

SR wrote the paper, analyzed data had primary responsibility for the final content. AS designed the research and analyzed data. RC performed data management and statistical analysis. MTR, DB and DKT designed the research. PDS analyzed data and wrote the paper. ZD designed the research, obtained the funding, and wrote the paper.

## References

1. Cani PD, Delzenne NM. The role of the gut microbiota in energy metabolism and metabolic disease. Curr Pharm Des. 2009;15(13):1546–1558. doi:10.2174/138161209788168164

2. Bidu C, Escoula Q, Bellenger S, et al. The Transplantation of ω3 PUFA-Altered Gut Microbiota of fat-1 Mice to Wild-Type Littermates Prevents Obesity and Associated Metabolic Disorders. Diabetes. 2018;67(8):1512–1523. doi:10.2337/db17-1488

3. Simopoulos AP. Evolutionary aspects of diet, the omega-6/omega-3 ratio and genetic variation: nutritional implications for chronic diseases. Biomed Pharmacother. 2006;60(9):502–507. doi:10.1016/j.biopha.2006.07.080

4. Bender N, Portmann M, Heg Z, Hofmann K, Zwahlen M, Egger M. Fish or n3-PUFA intake and body composition: a systematic review and meta-analysis. Obes Rev Off J Int Assoc Study Obes. 2014;15(8):657–665. doi:10.1111/obr.12189

5. Abbott KA, Burrows TL, Acharya S, Thota RN, Garg ML. DHA-enriched fish oil reduces insulin resistance in overweight and obese adults. Prostaglandins Leukot Essent Fatty Acids. 2020;159:102154. doi:10.1016/j.plefa.2020.102154

6. Wang J, Zhang Y, Zhao L. Omega-3 PUFA intake and the risk of digestive system cancers: A meta-analysis of observational studies. Medicine (Baltimore). 2020;99(19):e20119. doi:10.1097/MD.0000000000020119

7. Mani V, Hollis JH, Gabler NK. Dietary oil composition differentially modulates intestinal endotoxin transport and postprandial endotoxemia. Nutr Metab. 2013;10(1):6. doi:10.1186/1743-7075-10-6

8. Cani PD, Amar J, Iglesias MA, et al. Metabolic endotoxemia initiates obesity and insulin resistance. Diabetes. 2007;56(7):1761–1772. doi:10.2337/db06-1491

9. Caesar R, Tremaroli V, Kovatcheva-Datchary P, Cani PD, Bäckhed F. Crosstalk between Gut Microbiota and Dietary Lipids Aggravates WAT Inflammation through TLR Signaling. Cell Metab. 2015;22(4):658–668. doi:10.1016/j.cmet.2015.07.026

10. Monk JM, Liddle DM, Hutchinson AL, et al. Fish oil supplementation to a high-fat diet improves both intestinal health and the systemic obese phenotype. J Nutr Biochem. 2019;72:108216. doi:10.1016/j.jnutbio.2019.07.007

11. Durkin LA, Childs CE, Calder PC. Omega-3 Polyunsaturated Fatty Acids and the Intestinal Epithelium-A Review. Foods Basel Switz. 2021;10(1). doi:10.3390/foods10010199

12. Wilson MJ, Sen A, Bridges D, et al. Higher baseline expression of the PTGS2 gene and greater decreases in total colonic fatty acid content predict greater decreases in colonic prostaglandin-E2 concentrations after dietary supplementation with ω-3 fatty acids. Prostaglandins Leukot Essent Fatty Acids. 2018;139:14–19. doi:10.1016/j.plefa.2018.11.001

13. Bellenger J, Bellenger S, Escoula Q, Bidu C, Narce M. N-3 polyunsaturated fatty acids: An innovative strategy against obesity and related metabolic disorders, intestinal alteration and gut microbiota dysbiosis. Biochimie. 2019;159:66–71. doi:10.1016/j.biochi.2019.01.017

14. Kaliannan K, Wang B, Li X-Y, Kim K-J, Kang JX. A host-microbiome interaction mediates the opposing effects of omega-6 and omega-3 fatty acids on metabolic endotoxemia. Sci Rep. 2015;5:11276. doi:10.1038/srep11276

15. Pu S, Khazanehei H, Jones PJ, Khafipour E. Interactions between Obesity Status and Dietary Intake of Monounsaturated and Polyunsaturated Oils on Human Gut Microbiome Profiles in the Canola Oil Multicenter Intervention Trial (COMIT). Front Microbiol. 2016;7. doi:10.3389/fmicb.2016.01612

16. Everard A, Belzer C, Geurts L, et al. Cross-talk between Akkermansia muciniphila and intestinal epithelium controls diet-induced obesity. Proc Natl Acad Sci U S A. 2013;110(22):9066–9071. doi:10.1073/pnas.1219451110

17. Watson H, Mitra S, Croden FC, et al. A randomised trial of the effect of omega-3 polyunsaturated fatty acid supplements on the human intestinal microbiota. Gut. 2018;67(11):1974–1983. doi:10.1136/gutjnl-2017-314968

18. Ghosh S, DeCoffe D, Brown K, et al. Fish oil attenuates omega-6 polyunsaturated fatty acid-induced dysbiosis and infectious colitis but impairs LPS dephosphorylation activity causing sepsis. PloS One. 2013;8(2):e55468. doi:10.1371/journal.pone.0055468

19. Menni C, Zierer J, Pallister T, et al. Omega-3 fatty acids correlate with gut microbiome diversity and production of N-carbamylglutamate in middle aged and elderly women. Sci Rep. 2017;7(1):11079. doi:10.1038/s41598-017-10382-2

20. Flynn KJ, Ruffin MT, Turgeon DK, Schloss PD. Spatial Variation of the Native Colon Microbiota in Healthy Adults. Cancer Prev Res Phila Pa. 2018;11(7):393–402. doi:10.1158/1940-6207.CAPR-17-0370

21. Sidahmed E, Cornellier ML, Ren J, et al. Development of exchange lists for Mediterranean and Healthy Eating diets: implementation in an intervention trial. J Hum Nutr Diet Off J Br Diet Assoc. 2014;27(5):413–425. doi:10.1111/jhn.12158

22. Djuric Z, Bassis CM, Plegue MA, et al. Colonic Mucosal Bacteria Are Associated with Inter-Individual Variability in Serum Carotenoid Concentrations. J Acad Nutr Diet. 2018;118(4):606-616.e3. doi:10.1016/j.jand.2017.09.013

23. Kozich JJ, Westcott SL, Baxter NT, Highlander SK, Schloss PD. Development of a Dual-Index Sequencing Strategy and Curation Pipeline for Analyzing Amplicon Sequence Data on the MiSeq Illumina Sequencing Platform. Appl Environ Microbiol. 2013;79(17):5112–5120. doi:10.1128/AEM.01043-13

24. Pruesse E, Quast C, Knittel K, et al. SILVA: a comprehensive online resource for quality checked and aligned ribosomal RNA sequence data compatible with ARB. Nucleic Acids Res. 2007;35(21):7188–7196. doi:10.1093/nar/gkm864

25. Wang Q, Garrity GM, Tiedje JM, Cole JR. Naive Bayesian classifier for rapid assignment of rRNA sequences into the new bacterial taxonomy. Appl Environ Microbiol. 2007;73(16):5261–5267. doi:10.1128/AEM.00062-07

26. Westcott SL, Schloss PD. OptiClust, an Improved Method for Assigning Amplicon-Based Sequence Data to Operational Taxonomic Units. mSphere. 2017;2(2). doi:10.1128/mSphereDirect.00073-17

27. Noriega BS, Sanchez-Gonzalez MA, Salyakina D, Coffman J. Understanding the Impact of Omega-3 Rich Diet on the Gut Microbiota. Case Rep Med. 2016;2016:3089303. doi:10.1155/2016/3089303

28. Calder PC. Is Increasing Microbiota Diversity a Novel Anti-Inflammatory Action of Marine n-3 Fatty Acids? J Nutr. 2019;149(7):1102–1104. doi:10.1093/jn/nxz043

29. Costantini L, Molinari R, Farinon B, Merendino N. Impact of Omega-3 Fatty Acids on the Gut Microbiota. Int J Mol Sci. 2017;18(12). doi:10.3390/ijms18122645

30. Balfegó M, Canivell S, Hanzu FA, et al. Effects of sardine-enriched diet on metabolic control, inflammation and gut microbiota in drug-naïve patients with type 2 diabetes: a pilot randomized trial. Lipids Health Dis. 2016;15:78. doi:10.1186/s12944-016-0245-0

31. Backlund MG, Mann JR, Dubois RN. Mechanisms for the prevention of gastrointestinal cancer: the role of prostaglandin E2. Oncology. 2005;69 Suppl 1:28–32. doi:10.1159/000086629

32. Zheng J, Hoffman KL, Chen J-S, et al. Dietary inflammatory potential in relation to the gut microbiome: results from a cross-sectional study. Br J Nutr. Published online June 1, 2020:1-12. doi:10.1017/S0007114520001853

33. Tu P, Bian X, Chi L, et al. Characterization of the Functional Changes in Mouse Gut Microbiome Associated with Increased Akkermansia muciniphila Population Modulated by Dietary Black Raspberries. ACS Omega. 2018;3(9):10927–10937. doi:10.1021/acsomega.8b00064

34. Roopchand DE, Carmody RN, Kuhn P, et al. Dietary Polyphenols Promote Growth of the Gut Bacterium Akkermansia muciniphila and Attenuate High-Fat Diet-Induced Metabolic Syndrome. Diabetes. 2015;64(8):2847–2858. doi:10.2337/db14-1916

35. Jayachandran M, Chung SSM, Xu B. A critical review of the relationship between dietary components, the gut microbe Akkermansia muciniphila, and human health. Crit Rev Food Sci Nutr. 2020;60(13):2265–2276. doi:10.1080/10408398.2019.1632789

36. Anhê FF, Roy D, Pilon G, et al. A polyphenol-rich cranberry extract protects from diet-induced obesity, insulin resistance and intestinal inflammation in association with increased Akkermansia spp. population in the gut microbiota of mice. Gut. 2015;64(6):872–883. doi:10.1136/gutjnl-2014-307142

37. Willemsen LEM, Koetsier MA, van Deventer SJH, van Tol E a. F. Short chain fatty acids stimulate epithelial mucin 2 expression through differential effects on prostaglandin E(1) and E(2) production by intestinal myofibroblasts. Gut. 2003;52(10):1442–1447. doi:10.1136/gut.52.10.1442

38. Crittenden S, Goepp M, Pollock J, et al. Prostaglandin E2 promotes intestinal inflammation via inhibiting microbiota-dependent regulatory T cells. Sci Adv. 2021;7(7). doi:10.1126/sciadv.abd7954

39. Li Q, Zhang Q, Wang M, Zhao S, Xu G, Li J. n-3 polyunsaturated fatty acids prevent disruption of epithelial barrier function induced by proinflammatory cytokines. Mol Immunol. 2008;45(5):1356–1365. doi:10.1016/j.molimm.2007.09.003

40. Lallès J-P. Recent advances in intestinal alkaline phosphatase, inflammation, and nutrition. Nutr Rev. 2019;77(10):710–724. doi:10.1093/nutrit/nuz015

41. Sanguansri L, Shen Z, Weerakkody R, Barnes M, Lockett T, Augustin MA. Omega-3 fatty acids in ileal effluent after consuming different foods containing microencapsulated fish oil powder - an ileostomy study. Food Funct. 2013;4(1):74–82. doi:10.1039/c2fo30133d

42. Zhang Y-G, Xia Y, Lu R, Sun J. Inflammation and intestinal leakiness in older HIV+ individuals with fish oil treatment. Genes Dis. 2018;5(3):220–225. doi:10.1016/j.gendis.2018.07.001

